# Privacy-Preserving Multivariate Bayesian Regression Models for Overcoming Data Sharing Barriers in Health and Genomics

**DOI:** 10.1101/2025.07.30.25332448

**Authors:** Izel Fourie Sørensen, Peter Sørensen

## Abstract

We present multivariate Bayesian regression models specifically designed to overcome data-sharing barriers in health and genomics. These multi-response models are well suited for scenarios where data must remain decentralized due to privacy, intellectual property, or regulatory constraints. In extensive simulation studies, our approach consistently outperformed traditional single-response models trained on individual datasets, particularly under real-world conditions such as low signal, unbalanced cohorts, and high-dimensional feature spaces. For the first time, we demonstrate that multivariate Bayesian regression can be implemented using orthogonal transformations of sufficient statistics, enabling fully privacy-preserving analysis without sharing individual-level data. The models are scalable, interpretable, and applicable to predictive tasks across diverse collaborators, supporting secure data-driven research in domains such as clinical trials, biomarker discovery, and precision health.

## Introduction

In healthcare and biotechnology, enhanced predictions and insights are crucial for improving patient outcomes, identifying trends in population health, and reducing costs^1–3^. Achieving these benefits requires access to comprehensive datasets that enable development of more accurate predictive models. However, collecting additional data is often impractical or prohibitively expensive, making data sharing essential.

Despite its potential, data sharing faces significant challenges^4–7^. Stringent data privacy regulations, such as the General Data Protection Regulation (GDPR)^8^, limit the sharing of sensitive information, while technical issues, including the transfer of large datasets and heterogeneity among local datasets, complicate integration and analysis. Additionally, concerns over intellectual property rights and competitive interests discourage collaborators from sharing data. Overcoming these barriers is critical to advancing innovation in healthcare and beyond.

Federated learning (FL) has emerged as a promising strategy to support privacy-preserving collaboration^9,10^. FL enables decentralized model training across distributed datasets without requiring data to leave its source, thereby protecting privacy and minimizing data transfer. FL frameworks are increasingly applied in healthcare^7,11–16^, enabling predictive analytics and risk assessment across institutions while ensuring compliance with stringent privacy regulations like GDPR.

However, FL still faces several limitations. Data heterogeneity remains a key challenge, as local datasets often differ in size, quality, and distribution, leading to imbalanced contributions to global models^17,18^. In addition, communication and computation overhead can hinder scalability when working with large or high-dimensional datasets^19^. Finally, ensuring interpretability in FL-trained models remains an open problem, especially in sensitive domains like healthcare^20^.

To address these limitations, we propose a novel Bayesian federated learning framework based on Bayesian linear regression (BLR) models^21–26^. BLR provides a flexible foundation for statistical learning and prediction, accommodating both continuous and binary outcomes while handling correlated and high-dimensional predictors.

Our approach centers on a multivariate BLR model that leverages sufficient statistics to jointly analyze correlated outcomes^27–29^. This approach enables efficient computation, reduces complexity, and preserves key relationships in the data^28,30^. Though originally developed for genetic risk prediction^31,32^, we demonstrate that this framework generalizes well to other collaborative data-sharing scenarios. Importantly, our model uses only aggregated summary data, aligning with the privacy-preserving principles of federated learning. To further enhance privacy, we apply local orthogonal data transformations^33,34^ to encode sufficient statistics before transmission, ensuring that sensitive individual-level data remain fully protected. It is particularly suited for high-dimensional and heterogeneous data^28,29^, and its ability to incorporate prior knowledge and yield interpretable results enhances its practical utility.

In this study, we evaluate the performance of multivariate Bayesian linear regression (MT-BLR) models in federated learning settings. We compare two modeling strategies: (1) Single-response (ST) models, trained independently for each response variable and dataset, simulating scenarios where data are not shared across sources; and (2) Multi-response (MT) models, trained jointly across multiple response variables to estimate shared effects, allowing for information sharing across datasets. Both strategies were implemented within a Bayesian framework incorporating structured priors on predictor effects.

Our evaluation includes two components. First, we conduct an extensive simulation study across multiple scenarios to test model robustness under varying predictor densities, response correlations, and cohort size imbalances. Second, we apply the models to real-world data from the Social Diagnosis 2011 survey^35^ to illustrate their practical relevance. In both settings, we assess predictive performance and quantify how MT-BLR improves upon traditional single-response models, particularly when data are sparse or signal is shared.

## Methods

### Statistical models and analyses

#### Linear model

The foundation of the approach is a linear model expressed in matrix notation as:

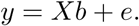

In this model, *y* represents the response variable, *X* is the design matrix of predictor variables, distributed with mean 0 and variance *σ*^2^. The vector *b* contains the regression coefficients corresponding to the predictor variables. The dimensions of *y*, *X*, *b* and *e* depend on the number of responses (*t*), predictor variables (*p*), and observations (*n*).

#### Single-response BLR model

A Bayesian linear regression (BLR) model with a spike-and-slab prior distribution^36^ was used to model the association between predictor variables and response variables for the single response BLR (ST-BLR) models:

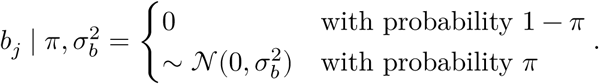

This assumes that the regression coefficients (*b*) are drawn from a mixture distribution consisting of a point mass at zero and a normal distribution with a common variance for all non-zero effects. Each coefficient *b_j_* is either zero or non-zero, where zero implies no contribution to the response variable, and a non-zero value indicates an association.

The prior inclusion probability (e.g., *π* = 0.001) specifies the expected proportion of non-zero regression coefficients. The prior distribution for the common variance of the non-zero coefficients follows an inverse chi-squared distribution, *χ*^−1^(*S_b_, v*_*b*_)^37^, where *S*_*b*_ is the scale parameter and *v_b_* is the degrees of freedom.

The mixture proportions are modeled using a Dirichlet distribution, Dirichlet(*C*, *c* +*α*), where *C* is the number of mixture components, *c* is the vector of counts of predictor variables assigned to each component, and *α* = (1, 1) is a concentration hyperparameter ensuring that the sampled proportions are governed by the observed data.

To support inference under this mixture model, a latent indicator vector *d* = (*d*_1_, *d*_2_, …, *d*_*m*_) is introduced via data augmentation, where each element *d_j_* indicates whether the *j*^th^ regression coefficient is zero or non-zero.

#### Multi-response BLR model

Bayesian linear regression models can be extended to accommodate multiple responses, which is useful for identifying shared effects of predictor variables across outcomes. A general multi-response BLR (MT-BLR) model based on the multivariate spike and slab prior distribution^36^ allows each predictor to influence any subset of responses, providing insight into whether predictors affect all, some, or none of the outcomes.

The MT-BLR model incorporates regularization similar to the single-response version while borrowing strength from correlations among regression coefficients across responses. In the case of two responses, the core regression equation for the regression coefficients can be expressed as:

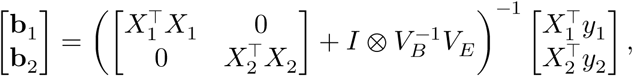

where ⊗ denotes the Kronecker product. In this model, the key parameters include the covari-capture matrix of the regression coefficients, *V_B_*, and the residual covariance matrix, *V_E_*, which capture dependencies among effects and residuals across responses.

For the two-response case, the regression coefficient covariance matrix *V_B_* is:

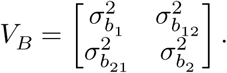

If *V_B_* is not assumed constant across predictor variables, the model allows for differential shrinkage of regression coefficients, which can be incorporated using spike-and-slab priors. Moreover, if the covariance between coefficients for different responses (e.g., 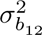) is non-zero, the model can borrow information across responses, increasing power to detect associations.

The residual covariance matrix *V_E_* is similarly defined as:

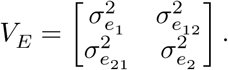

This matrix captures residual variation and correlation not explained by the regression coefficients, including response-specific variability and measurement error.

The prior distribution for the regression coefficient covariance matrix is 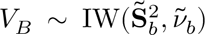, residual where *ṽ_b_* is the degrees of freedom and 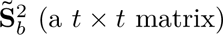 is the scale matrix. Similarly, the residual covariance matrix is 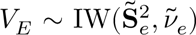, where *ṽ_e_* is the degrees of freedom and 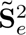 is the corresponding scale matrix.

#### Extension to sufficient statistics

The Bayesian regression model is implemented using sufficient statistics rather than individual-level data. Specifically, the algorithm requires only the cross-product matrices of the design matrix and the response vector, *X*^⊤^ *X* and *X*^⊤^*y*, which are sufficient for estimating regression parameters under Gaussian likelihood assumptions.

This formulation enables the model to operate directly on pre-computed summary statistics, making it well suited for settings where individual-level data are unavailable or restricted due to privacy or data-sharing constraints. By avoiding direct access to raw data, the method supports scalable and privacy-preserving analyses in large-scale applications such as genomic studies, electronic health records, and other biomedical domains, where legal, ethical, or logistical barriers often hinder data sharing.

#### Privacy-aware summary statistic sharing

To enhance privacy while enabling multivariate Bayesian regression, each collaborating site provides only rotated sufficient statistics derived from local data. Specifically, an eigen-decomposition is performed on the local cross-product matrix, *X*^⊤^*X* = *U* Λ*U*^⊤^, and the sufficient statistics are rotated by multiplying with the scaled eigenvectors, Λ^−0.5^*U*^⊤^. Only these rotated sufficient statistics are shared across sites, while the original data and the decomposition matrices (*U*, Λ) remain local.

This transformation preserves the statistical integrity of the model while introducing a practical layer of privacy protection. Because the transformation is not reversible without access to the local eigenvectors, it enables secure, privacy-aware computation without compromising inference accuracy.

#### Estimation of BLR model parameters

Parameter estimates for the BLR model (e.g., *b_j_, π*, 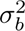, and 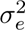 for the single-response model) were obtained using Markov Chain Monte Carlo (MCMC) via Gibbs sampling. Further details on the procedure are provided by Rohde et al.^28^. For both ST and MT analyses, 3000 iterations were used, with the first 500 discarded as burn-in. Multiple chains were run to ensure convergence.

### Evaluation of Bayesian models in simulated data-sharing scenarios

To evaluate the predictive performance of Bayesian regression models in multi-dataset settings, we applied both ST and MT approaches to a series of simulated datasets. Each simulation replicate consisted of response variables from three related datasets and a shared predictor matrix. Depending on the scenario, the training sample size varied across datasets to mimic both balanced and imbalanced data-sharing conditions.

Response variables were standardized and partitioned into training and test sets. Summary statistics, including cross-product matrices of predictors and outcome vectors, were computed separately for each dataset. ST models were fit independently using Bayesian linear regression with a BayesC prior. Prediction accuracy was assessed using the coefficient of determination (R²).

In parallel, we evaluated four MT-BLR models:

- MT1: Joint model assuming shared predictor effects across datasets
- MT2: Restrictive variant of MT1 with constrained sharing
- MT3: Incorporates known groupings of informative and uninformative predictors
- MT4: Restrictive version of MT3 with structured prior assumptions

All MT models were fit jointly across datasets using the same prior as the ST model. Accuracy was calculated per dataset and replicate, and improvements over ST-BLR were quantified as the percentage change in R².

Model performance was assessed across 48 simulated scenarios that varied along four key factors: the proportion of variance explained (PVE), set at 0.05 or 0.10; the number of predictors (P), set at 100 or 1000; the correlation between responses (*ρ*), set at 0.5 (representing heterogeneous effects) or 1.0 (homogeneous effects); and the training sample size configuration, which included equal-sized datasets (500×3, 1000×3, or 2000×3) as well as unequal-sized datasets (500/1000/2000, 500/500/1000, and 750/750/1000).

These scenarios reflect diverse multi-dataset prediction challenges, including balanced vs. imbalanced sample sizes, sparse vs. dense signal settings, and varying levels of correlations across response variables.

### Data-sharing models applied to survey data

In order to further demonstrate the applicability of the data-sharing models, they were applied to real-world data. We analyzed data from the Social Diagnosis 2011 (SD2011) survey, a publicly available dataset included in the R package synthpop^35^. SD2011 is derived from the Polish panel survey Conditions and Quality of Life of the Polish Population and includes responses from 5000 individuals on a wide range of demographic, socioeconomic, and health-related variables.

For this study, we prepared a cleaned dataset as follows: variables with more than 700 missing values were excluded, and all remaining records containing missing data were removed. Individuals reporting negative income values were also excluded. The marital status variable was recoded to group all forms of separation under a single “separated” category. Additionally, variables deemed redundant or requiring further transformation (*wkabint, workab, ls, trustfam, wkabdur, alcsol*, and *eduspec*) were removed. The resulting dataset, consisting of 3355 observations and 29 variables, was used for all analyses, with *income* as the continuous response variable.

To simulate federated data scenarios, the dataset was repeatedly partitioned into training and test sets using an 80/20 stratified split, repeated over 20 replicates. The training data were distributed across three virtual collaborators using two different partitioning strategies. In the first scenario, the training data were divided in a 1:2:3 ratio, resulting in unequal collaborator sample sizes of 448, 896, and 1342. In the second scenario, the training data were evenly distributed across the three collaborators, yielding approximately equal sample sizes of 896, 895, and 895.

Within each collaborator, summary statistics were computed and used to fit BLR models with BayesC priors. Models were trained using 3000 MCMC iterations with a burn-in of 500. In addition to single-response (ST) models, we evaluated two multi-response (MT) models. MT1 is a joint model assuming shared predictor effects across collaborators, while MT2 is a restrictive variant of MT1 that limits the extent of sharing.

All models were evaluated on the held-out test set. Predictive performance was assessed using R², from regressing observed outcomes on predicted values. Results were averaged across replicates, and improvements over ST were quantified as the percentage change in R².

## Results

### Evaluation of Bayesian models in simulated data-sharing scenarios

We compared the relative prediction accuracy of multi-response Bayesian models (MT1–MT4) to the single-response (ST) baseline under equal and unequal training set size conditions (Figure 1). In the equal training size scenarios (e.g., 500×3 and 2000×3, Figure 1 panels A and B), all MT models consistently outperformed the ST model across datasets, particularly when the proportion of variance explained (PVE) and the number of predictors (P) were higher. MT4 yielded the largest improvements, reflecting the advantage of jointly modeling related datasets with partially shared structure.

**Figure 1:**
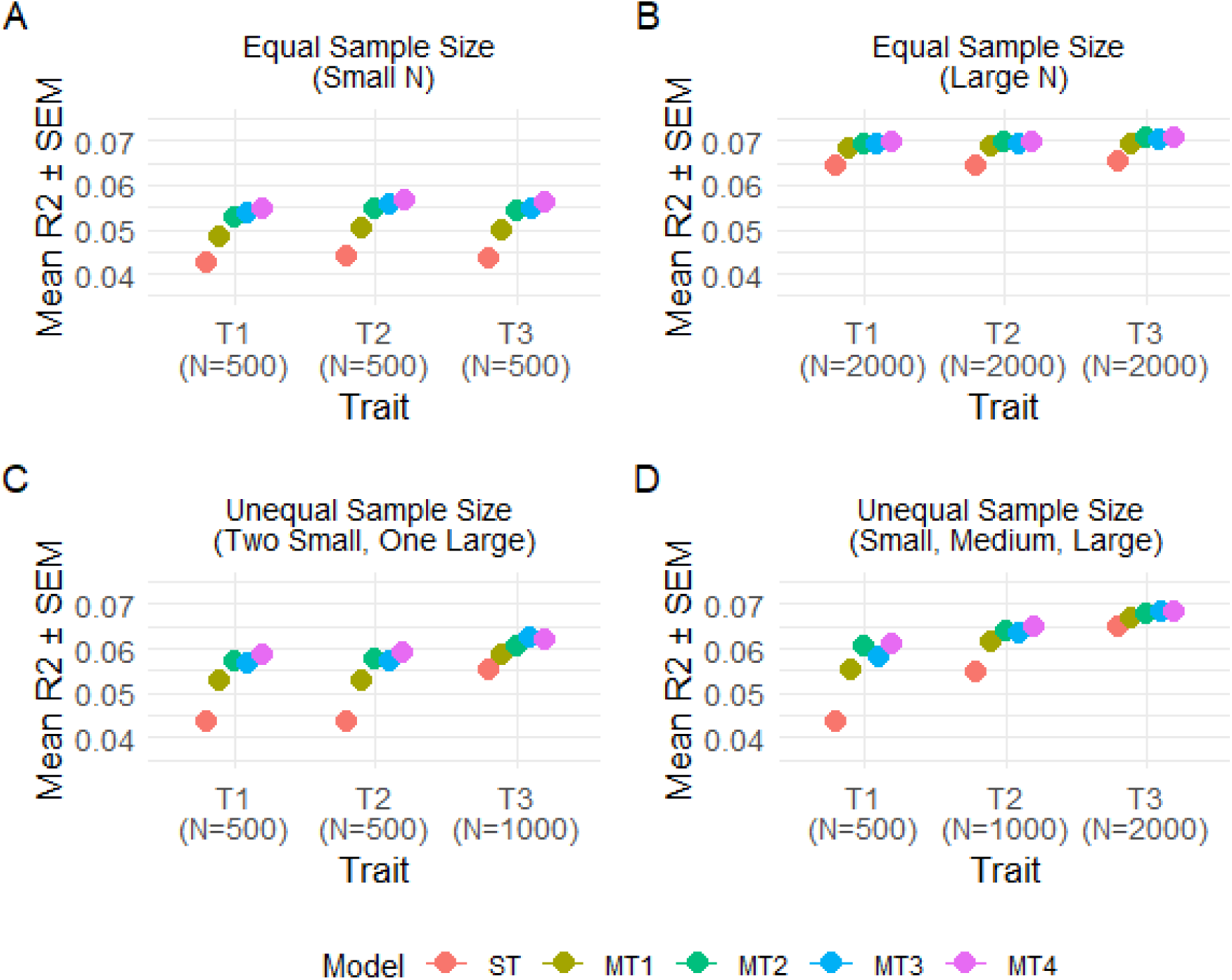
Prediction accuracy (mean ± SEM of R²) across responses and models under four training sample size configurations. Each panel shows results for a different config-uration: (A) equal sample size with small N (500 per response), (B) equal sample size with large N (2000 per response), (C) unequal sample sizes with two small (500) and one large (1000), and (D) unequal sample sizes increasing from small (500) to medium (1000) to large (2000). Prediction accuracy is plotted for each response (T1–T3), with models distinguished by color. Results are averaged over replicates.

In the unequal training size scenarios (e.g., 500/500/1000, Figure 1 panel C), performance gains from MT models were preserved but slightly reduced, particularly for datasets with larger training samples. These results indicate that while MT models are robust to imbalanced training sizes, the predictive benefit is greatest when all datasets are well represented.

We further evaluated prediction accuracy across key simulation conditions (Figure 2).

**Figure 2:**
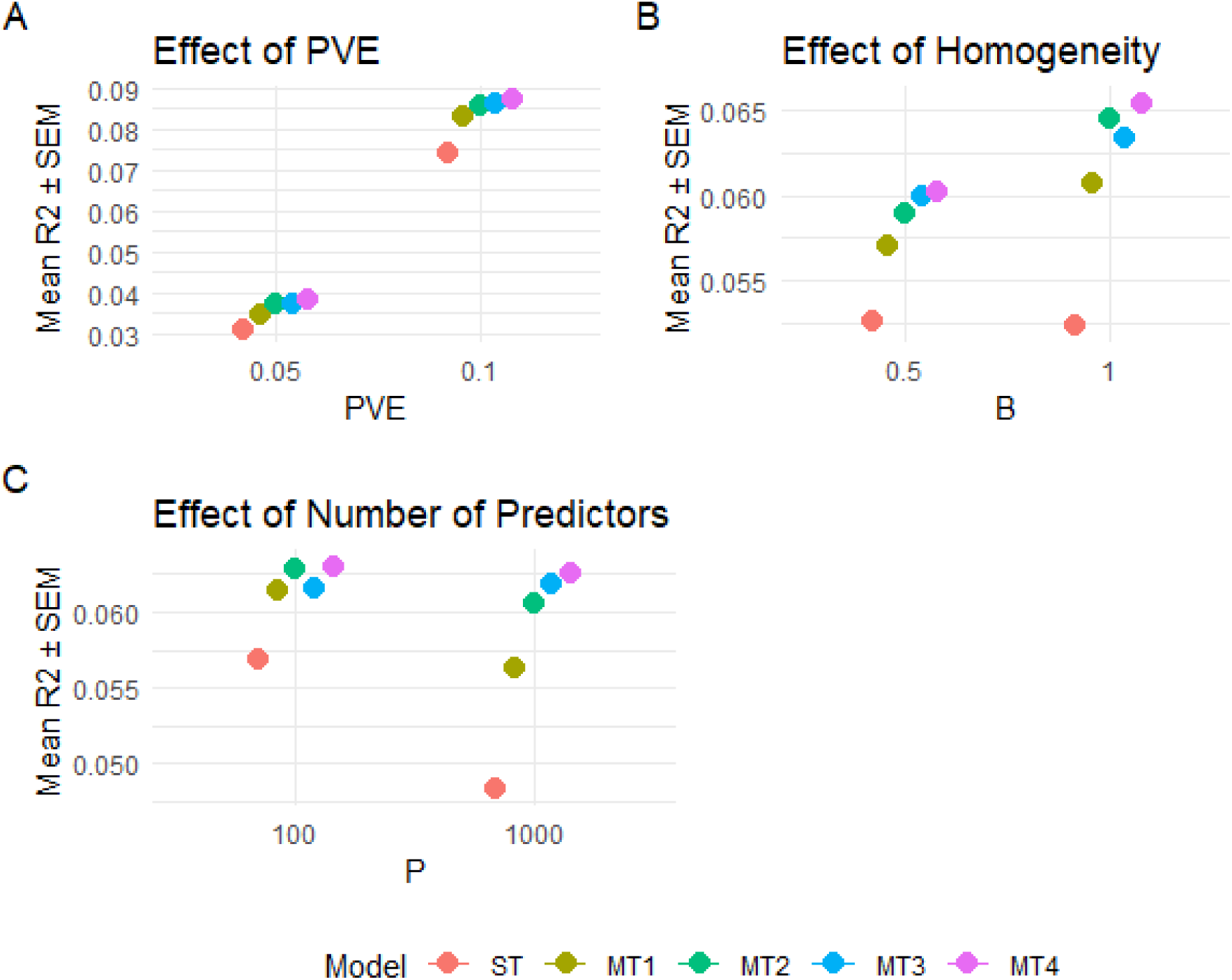
Comparison of prediction performance (mean R² ± SEM) for each model across key simulation parameters: (A) impact of PVE, (B) effect of predictor effect homogeneity (HET: B = 0.5, HOM: B = 1) and (C) influence of number of predictors (P). All results are averaged over replicates. Error bars show the standard error of the mean. Model ordering is consistent across panels.

When signal strength was low (PVE = 0.05), the ST model achieved a mean R² of 0.030, while MT models showed increasing gains: MT1 (0.033) to MT4 (0.041), with MT4 improving ∼38% over ST. At higher signal strength (PVE = 0.10), all models improved, with ST reaching 0.070 and MT4 0.090 (∼28% gain), underscoring the benefits of information sharing when the signal-to-noise ratio is low.

We also assessed the effect of dataset similarity. When predictor effects were only partially shared across datasets (B = 0.5), MT4 improved ∼25% over ST. Under full sharing (B = 1.0), the gains were larger with MT4 achieving an R² of 0.069 versus 0.050 for ST (∼38% gain). These results confirm that multi-response models are effective under both moderate and strong cross-dataset correlation.

Finally, in high-dimensional settings (P = 1000), prediction accuracy dropped for the ST model (R² = 0.044), but MT models maintained robust performance, with MT4 achieving 0.066 (∼49% improvement). This deomstrates the scalability of MT models in challenging, high-dimensional prediction tasks.

### Analysis of Social Diagnosis survey data

We applied BLR models to predict *income* based on selected variables from the Social Diagnosis 2011 dataset, evaluating the performance of multi-response approaches under varying dataset size conditions.

Across both equal and unequal dataset size settings (Figure 3), multi-response models (MT1 and MT2) consistently outperformed the single-response (ST) model in terms of predictive accuracy. When training datasets were of equal size (Figure 3, Panels A and C), MT2 achieved the highest mean R² values across datasets (0.290–0.291), followed closely by MT1 (0.286– 0.288), while the ST model performed worst (0.272–0.275). These differences translated into statistically significant improvements in prediction accuracy for both MT1 and MT2 relative to ST (p < 0.001).

**Figure 3:**
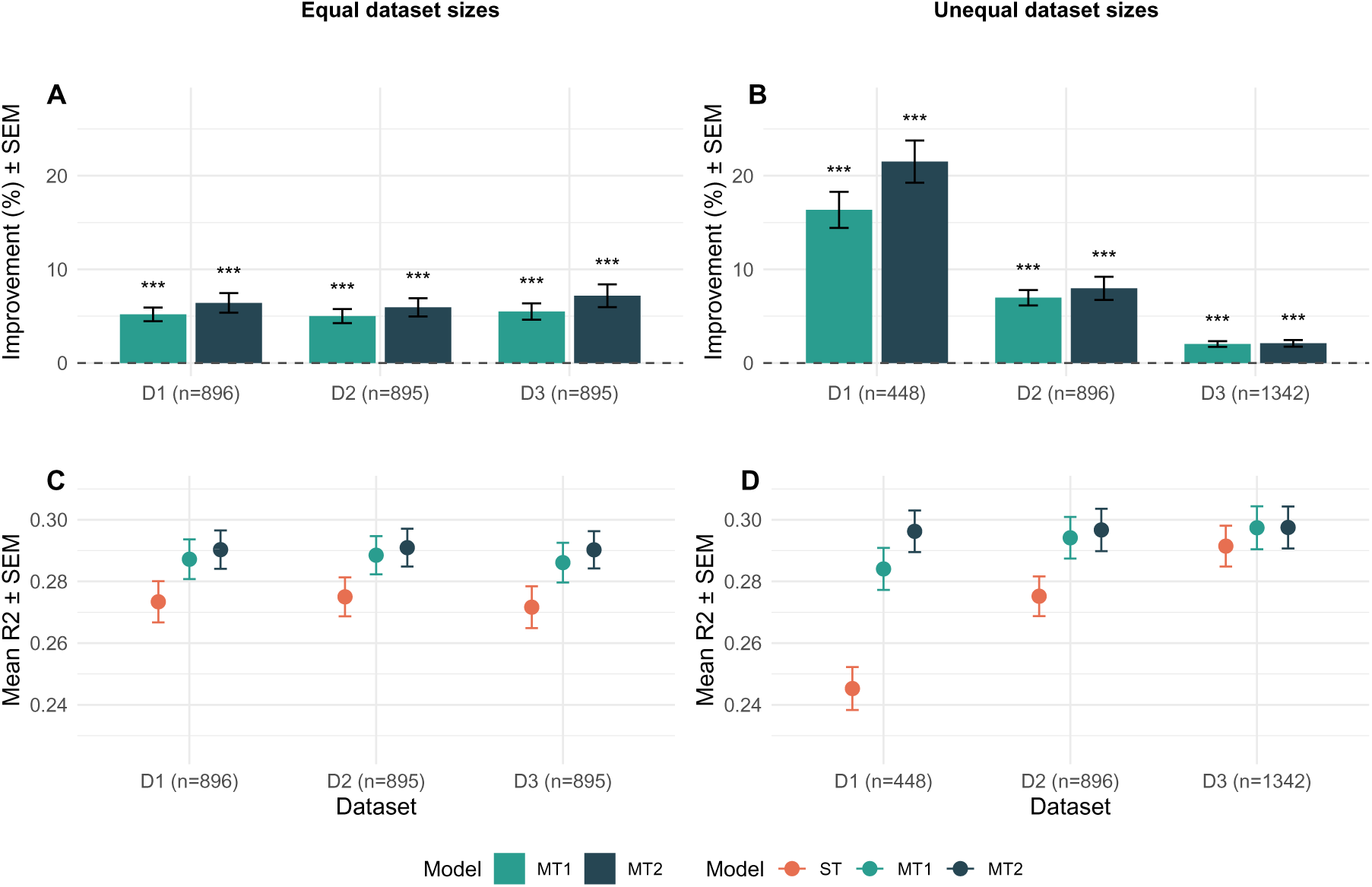
Comparison of prediction accuracy across datasets and modeling approaches. Each panel shows results from Bayesian linear regression applied to synthetic responses derived from the Social Diagnosis 2011 data. Panels A and B display the mean percentage improvement in prediction accuracy relative to a single-response model (ST) for multi-response models MT1 (flexible) and MT2 (restrictive), with error bars representing the standard error of the mean. Asterisks indicate statistical significance of improvement based on a one-sample t-test against zero. In all cases, improvements were significant at p < 0.001 (***). Panels C and D show the corresponding mean R² values (± SEM) for each model. Results are presented separately for settings with equal dataset sizes (A, C) and unequal dataset sizes (B, D).

The benefit of multi-response modeling was even more pronounced in the unequal dataset size setting (Figure 3, Panels B and D), particularly for the smallest dataset (D1, n = 448). Here, MT2 showed a substantial gain, reaching a mean R² of 0.296 compared to 0.284 for MT1 and only 0.245 for ST. The corresponding improvement exceeded 20% for MT2 and remained significant (p < 0.001). As dataset size increased (datasets D2 and D3 in Figure 3), differences in R² narrowed across models, indicating that the advantage of multi-response approaches diminishes with larger sample sizes but remains consistent. These findings underscore the utility of multi-response modeling, especially MT2, for improving prediction accuracy, particularly when individual-level data are limited.

## Discussion

In this study, we introduced multivariate Bayesian linear regression models designed to enable collaborative analysis under strict data-sharing constraints. These models are well suited for use cases where individual-level data must remain local due to privacy, regulatory, or competitive concerns. Through extensive simulation and real-world applications, we showed that our approach outperforms traditional single-response (ST) models, particularly in settings with low signal-to-noise ratios, unbalanced datasets, and high-dimensional predictors. Importantly, we show for the first time that multivariate Bayesian regression can be implemented using local orthogonal data transformations, enabling fully privacy-preserving inference without requiring access to raw data. This approach aligns closely with federated learning principles, allowing collaborators to train shared models without pooling sensitive information. The resulting framework is scalable, interpretable, and deployable across domains such as clinical trials, biomarker discovery, and precision health.

### Insights from simulation study

Our simulation results show that multi-response models can substantially improve predictive performance compared to single-response models across a wide range of scenarios. These included both low and high signal conditions, varying degrees of similarity between datasets, and unequal training set sizes. Across all settings, MT-BLR consistently outperformed the single-response baseline. Among the model variants, MT4, which incorporates prior knowledge about predictor groupings, delivered the most consistent gains in prediction accuracy as measured by R².

These findings highlight the flexibility of MT-BLR for collaborative data-sharing applications. The framework is suitable for analyses within a single institution that collects multiple outcomes under different conditions, as well as for multi-institutional studies where datasets may differ in outcomes, covariates, or both. Whether collaborators work with identical outcomes and predictors, partially overlapping features, or entirely distinct datasets, MT-BLR can effectively borrow information to improve prediction.

In addition to improved accuracy, the framework offers interpretable outputs, including R², AUC, variable inclusion probabilities, and importance scores. These summaries help identify the most influential predictors and clarify how each dataset contributes to the global model.

The modeling strategy integrates modern Bayesian machine learning techniques that support a wide range of data environments. It performs well with both small and large sample sizes, handles high-dimensional predictor spaces, and enables joint modeling of continuous and binary outcomes. Built-in tools for variable selection and regularization help ensure model stability and interpretability.

Together, these results confirm that MT-BLR is robust and adaptable to a variety of collaborative data scenarios. The joint modeling approach provides substantial value, especially in cases where the signal is weak or where individual datasets are noisy or limited.

### Application to real data

The utility of MT-BLR models was further demonstrated in the analysis of the real-world Social Diagnosis survey data, where both MT1 and MT2 outperformed the ST baseline in predicting income. Improvements were most pronounced in settings with limited training data, demonstrating that MT-BLR can leverage information from larger, related datasets to improve prediction for smaller partners. These results reinforce the practical utility of MT-BLR in real-world data-sharing environments.

### Privacy preserving approach

We introduce a novel approach to privacy-preserving analysis using local orthogonal transformations of sufficient statistics. Each collaborator rotates their summary data using a private transformation matrix before transmission. These rotations obscure the original data while preserving the information needed for valid Bayesian inference. Since rotation matrices are kept private, this acts as a secure encoding step.

This design aligns conceptually with federated learning and secure multi-party computation, allowing effective model training without centralized data pooling. To our knowledge, this is the first implementation of a multivariate BLR framework using locally rotated sufficient statistics for secure, collaborative analysis. Future work should further investigate the theoretical privacy guarantees of this transformation, as well as its empirical performance in more heterogeneous and high-stakes real-world settings.

### Implementation

We believe our approach is readily implementable in practice, as illustrated in Figure 4. Each collaborator locally transforms and encrypts their summary statistics, which are securely transmitted to a central platform. There, they are aggregated to train a global model, which is returned to each partner for local evaluation. Partners can assess the predictive performance of the global model on their own data and extract insights, all while keeping individual-level data local. This enables knowledge sharing without compromising data privacy.

**Figure 4:**
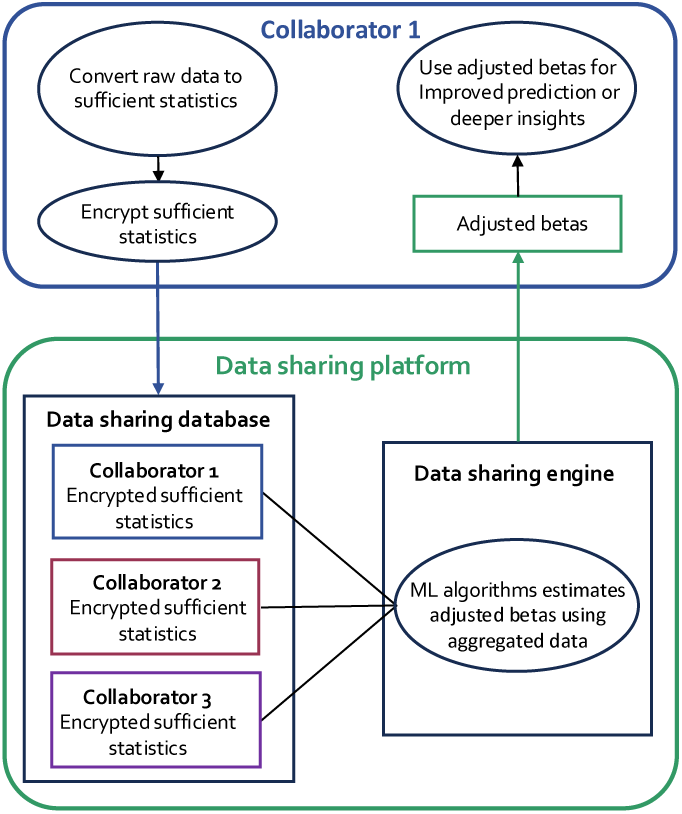
A schematic presentation of how our data-sharing method with multivariate Bayesian regression models are implemented. Each collaborator converts their raw data into sufficient statistics followed by encryption of the data. These encrypted summary statistics are sent to a secure data-sharing platform, where they are aggregated. Multivariate Bayesian regression models are then trained on the aggregated data to generate a global model that incorporates shared insights. The global model is returned to each collaborator, who applies it to generate more accurate predictions and derive deeper insights.

Importantly, our framework reveals how much each dataset contributes to the global model and highlights variables with shared versus divergent signals. These diagnostic tools can detect inconsistent effects, enhance transparency, and guide collaborative model refinement.

We provide an open-source R package to support adoption.

### Limitations

There are some limitations to our study. First, the current investigation assumes no missing data. While this simplifies analysis, missing covariates can be addressed using mean or median imputation, k-nearest neighbors, or Bayesian methods^38^. For missing values in response variables, we have previously shown that multivariate Bayesian regression is robust to small-to-moderate levels of missingness^29^. Alternatively, missing responses can be imputed during each iteration of the Gibbs sampler by drawing from their full conditional distributions^37^. Nevertheless, handling arbitrary missing data patterns in multivariate models remains a complex challenge and warrants further research.

Second, our current implementation focuses on multivariate regression with continuous outcomes. While this is a common case in biomedical and social science applications, extensions to other outcome types such as binary, ordered categorical, and right-censored Gaussian outcomes are possible using existing Bayesian methods, including those described by Korsgaard et al.^39^ and Yang et al.^40^. Future work should explore these extensions to broaden the applicability of our framework.

Third, we have not yet compared our method to foundation models or large pre-trained architectures. Although these models have achieved strong results on large, homogeneous datasets, they often underperform in low-resource settings or when datasets differ in covariate structure, phenotype distribution, or measurement protocols. In contrast, our approach is specifically designed to remain effective under such heterogeneous conditions. Recent studies have underscored the limitations of foundation models in these settings^41,42^. A systematic comparison between MT-BLR and foundation models in cross-site, small-sample environments represents an important direction for future research.

## Conclusion

MT-BLR offers a practical, interpretable, and privacy-preserving solution for collaborative modeling. It overcomes key limitations of standard federated learning by combining multivariate Bayesian modeling with local data transformations. Our approach enables secure, scalable, and accurate prediction across distributed datasets where data sharing is restricted.

## Data Availability

All data produced in the present study are available upon reasonable request to the authors.

## Appendix

**Figure 5:**
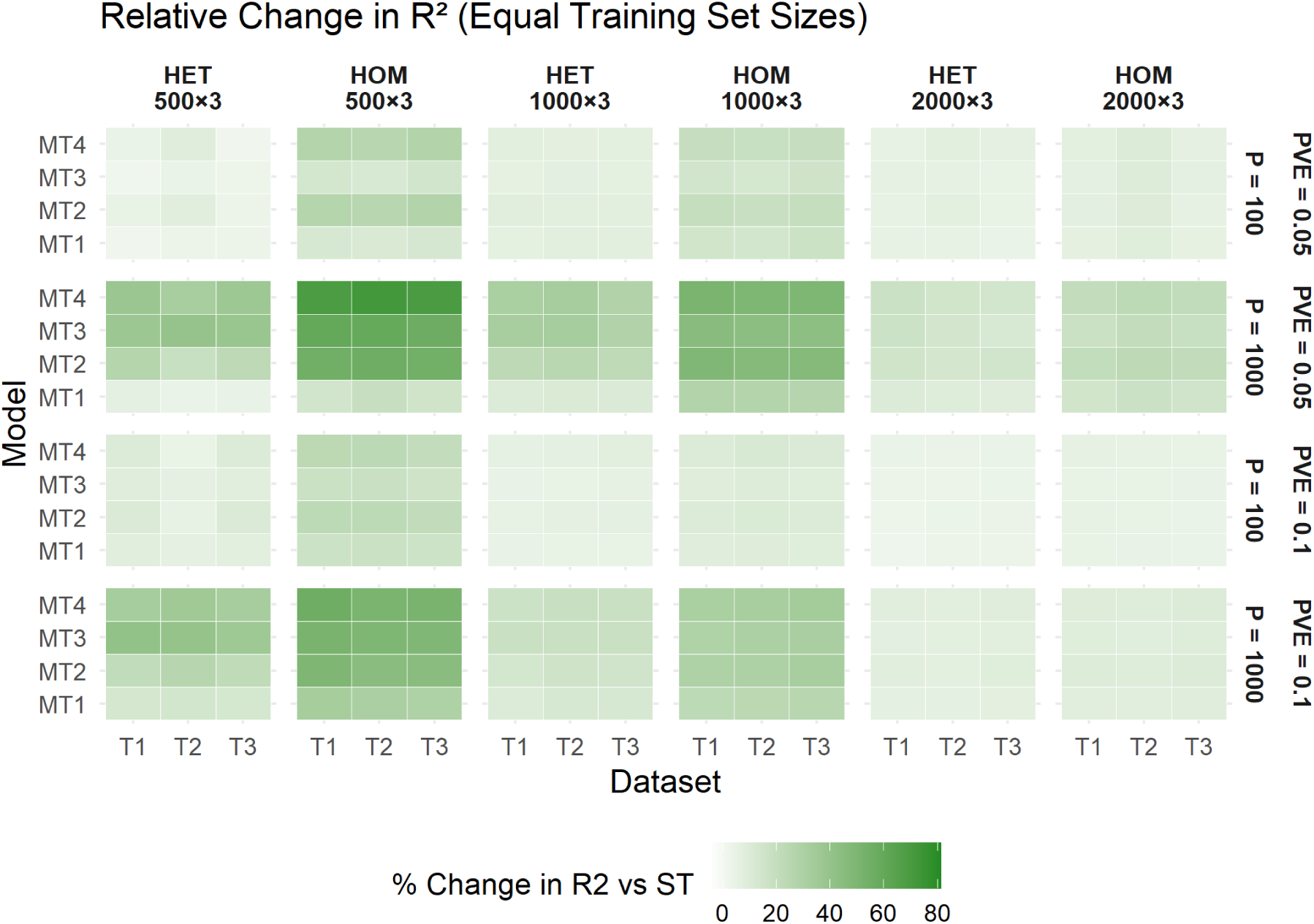
Relative improvement in prediction accuracy (R²) for multi-response Bayesian models (MT1–MT4) compared to the single-response baseline (ST), under scenarios with equal training set sizes (e.g., 1000×3). Each tile represents the percent difference in R² between an MT model and ST for a given dataset. Facets indicate the training configuration and degree of predictor effect similarity across datasets (HOM vs. HET), stratified by the proportion of variance explained (PVE) and number of predictors (P). Green tiles reflect improved performance relative to ST.

**Figure 6:**
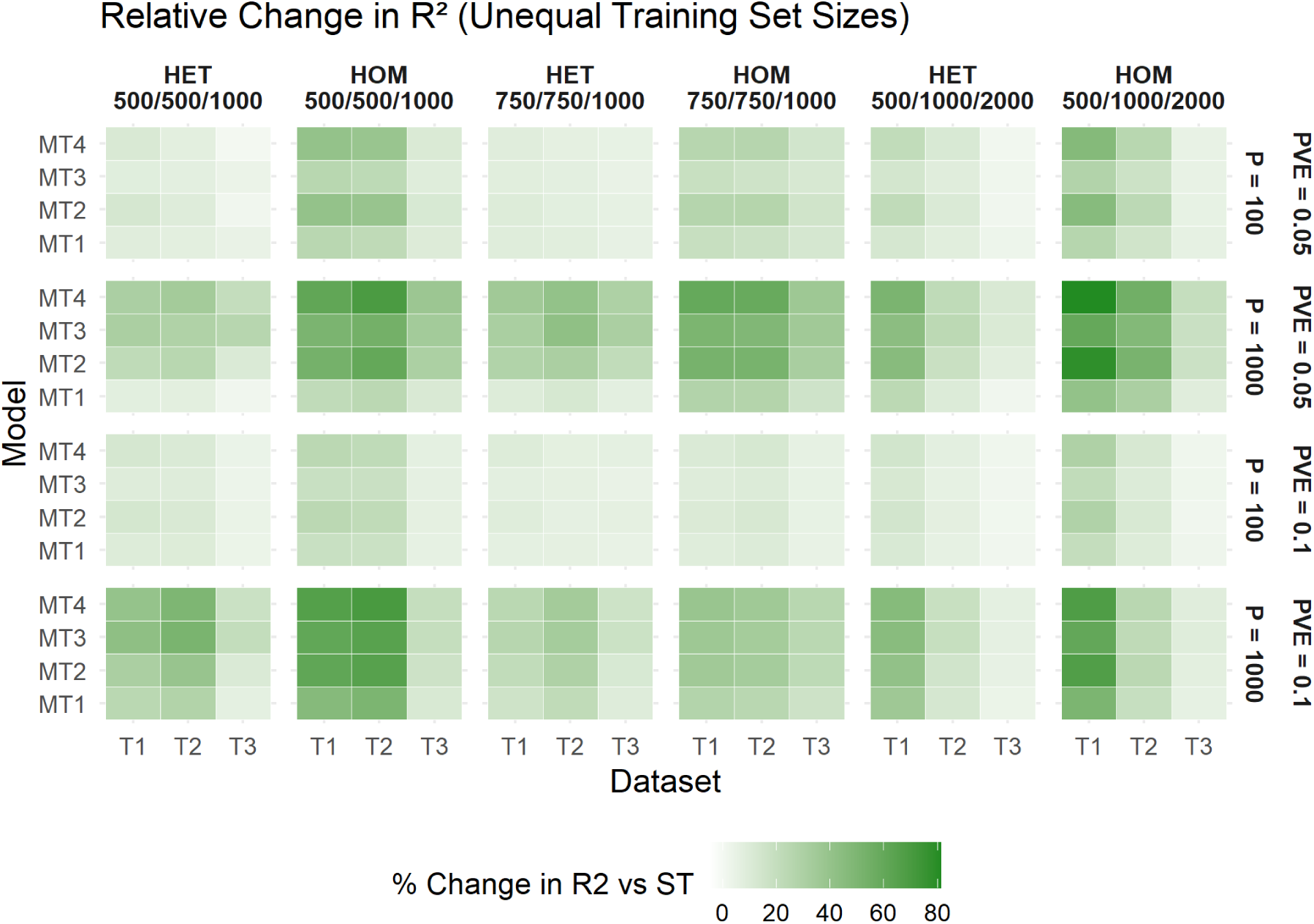
Relative improvement in prediction accuracy (R²) for multi-response Bayesian models (MT1–MT4) compared to the single-response baseline (ST), under scenarios with unequal training set sizes (e.g., 500/500/1000). Each tile represents the percent difference in R² between an MT model and ST for a given dataset. Facets indicate the training configuration and degree of predictor effect similarity across datasets (HOM vs. HET), stratified by the proportion of variance explained (PVE) and number of predictors (P). Green tiles reflect improved performance relative to ST.

**Table 1:** Overview of simulation scenarios with varying proportion of variance explained (PVE), number of predictors (P), correlation structure, and training set sizes.

| Scenario | PVE | P | Correlation | Training Set Sizes |
| --- | --- | --- | --- | --- |
| sim1 | 0.05 | 100 | 0.5 | 500×3 |
| sim2 | 0.10 | 100 | 0.5 | 500×3 |
| sim3 | 0.05 | 1000 | 0.5 | 500×3 |
| sim4 | 0.10 | 1000 | 0.5 | 500×3 |
| sim5 | 0.05 | 100 | 1.0 | 500×3 |
| sim6 | 0.10 | 100 | 1.0 | 500×3 |
| sim7 | 0.05 | 1000 | 1.0 | 500×3 |
| sim8 | 0.10 | 1000 | 1.0 | 500×3 |
| sim9 | 0.05 | 100 | 0.5 | 1000×3 |
| sim10 | 0.10 | 100 | 0.5 | 1000×3 |
| sim11 | 0.05 | 1000 | 0.5 | 1000×3 |
| sim12 | 0.10 | 1000 | 0.5 | 1000×3 |
| sim13 | 0.05 | 100 | 1.0 | 1000×3 |
| sim14 | 0.10 | 100 | 1.0 | 1000×3 |
| sim15 | 0.05 | 1000 | 1.0 | 1000×3 |
| sim16 | 0.10 | 1000 | 1.0 | 1000×3 |
| sim17 | 0.05 | 100 | 0.5 | 2000×3 |
| sim18 | 0.10 | 100 | 0.5 | 2000×3 |
| sim19 | 0.05 | 1000 | 0.5 | 2000×3 |
| sim20 | 0.10 | 1000 | 0.5 | 2000×3 |
| sim21 | 0.05 | 100 | 1.0 | 2000×3 |
| sim22 | 0.10 | 100 | 1.0 | 2000×3 |
| sim23 | 0.05 | 1000 | 1.0 | 2000×3 |
| sim24 | 0.10 | 1000 | 1.0 | 2000×3 |
| sim25 | 0.05 | 100 | 0.5 | 500/1000/2000 |
| sim26 | 0.10 | 100 | 0.5 | 500/1000/2000 |
| sim27 | 0.05 | 1000 | 0.5 | 500/1000/2000 |
| sim28 | 0.10 | 1000 | 0.5 | 500/1000/2000 |
| sim29 | 0.05 | 100 | 1.0 | 500/1000/2000 |
| sim30 | 0.10 | 100 | 1.0 | 500/1000/2000 |
| sim31 | 0.05 | 1000 | 1.0 | 500/1000/2000 |
| sim32 | 0.10 | 1000 | 1.0 | 500/1000/2000 |
| sim33 | 0.05 | 100 | 0.5 | 500/500/1000 |
| sim34 | 0.10 | 100 | 0.5 | 500/500/1000 |
| sim35 | 0.05 | 1000 | 0.5 | 500/500/1000 |
| sim36 | 0.10 | 1000 | 0.5 | 500/500/1000 |
| sim37 | 0.05 | 100 | 1.0 | 500/500/1000 |
| sim38 | 0.10 | 100 | 1.0 | 500/500/1000 |
| sim39 | 0.05 | 1000 | 1.0 | 500/500/1000 |
| sim40 | 0.10 | 1000 | 1.0 | 500/500/1000 |
| sim41 | 0.05 | 100 | 0.5 | 750/750/1000 |
| sim42 | 0.10 | 100 | 0.5 | 750/750/1000 |
| sim43 | 0.05 | 1000 | 0.5 | 750/750/1000 |
| sim44 | 0.10 | 1000 | 0.5 | 750/750/1000 |
| sim45 | 0.05 | 100 | 1.0 | 750/750/1000 |
| sim46 | 0.10 | 100 | 1.0 | 750/750/1000 |
| sim47 | 0.05 | 1000 | 1.0 | 750/750/1000 |
| sim48 | 0.10 | 1000 | 1.0 | 750/750/1000 |

**Table 2:**
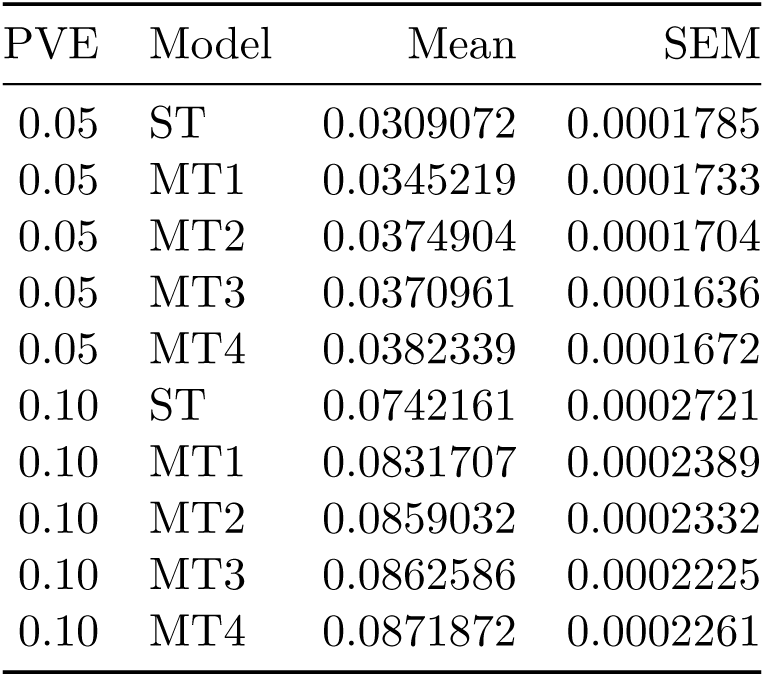
Effect of Proportion of Variance Explained (PVE) on R².

**Table 3:** Effect of Correlation Between Effects Across Datasets (B) on R².

| B | Model | Mean | SEM |
| --- | --- | --- | --- |
| 0.5 | ST | 0.0526832 | 0.0003440 |
| 0.5 | MT1 | 0.0570062 | 0.0003547 |
| 0.5 | MT2 | 0.0589582 | 0.0003527 |
| 0.5 | MT3 | 0.0599715 | 0.0003485 |
| 0.5 | MT4 | 0.0601564 | 0.0003522 |
| 1.0 | ST | 0.0524401 | 0.0003433 |
| 1.0 | MT1 | 0.0606864 | 0.0003532 |
| 1.0 | MT2 | 0.0644354 | 0.0003460 |
| 1.0 | MT3 | 0.0633832 | 0.0003492 |
| 1.0 | MT4 | 0.0652647 | 0.0003460 |

**Table 4:** Effect of Number of Predictors (P) on R².

| P | Model | Mean | SEM |
| --- | --- | --- | --- |
| 100 | ST | 0.0568631 | 0.0003412 |
| 100 | MT1 | 0.0614704 | 0.0003484 |
| 100 | MT2 | 0.0628643 | 0.0003483 |
| 100 | MT3 | 0.0615508 | 0.0003486 |
| 100 | MT4 | 0.0629333 | 0.0003498 |
| 1000 | ST | 0.0482603 | 0.0003385 |
| 1000 | MT1 | 0.0562222 | 0.0003580 |
| 1000 | MT2 | 0.0605293 | 0.0003529 |
| 1000 | MT3 | 0.0618039 | 0.0003502 |
| 1000 | MT4 | 0.0624878 | 0.0003510 |

